# Modelling the effectiveness and social costs of daily lateral flow antigen tests versus quarantine in preventing onward transmission of COVID-19 from traced contacts

**DOI:** 10.1101/2021.08.06.21261725

**Authors:** Luca Ferretti, Chris Wymant, Anel Nurtay, Lele Zhao, Robert Hinch, David Bonsall, Michelle Kendall, Joanna Masel, John Bell, Susan Hopkins, A. Marm Kilpatrick, Tim Peto, Lucie Abeler-Dörner, Christophe Fraser

**Affiliations:** Big Data Institute, Li Ka Shing Centre for Health Information and Discovery, Nuffield Department of Medicine, University of Oxford, Oxford, UK; Wellcome Centre for Human Genetics, Nuffield Department of Medicine, University of Oxford, Oxford, UK; Department of Statistics, University of Warwick, Coventry, UK; Department of Ecology and Evolutionary Biology, University of Arizona, Tucson AZ, USA; WeHealth Solutions PBC, Sacramento CA, USA; Nuffield Department of Medicine, University of Oxford, Oxford, UK; National Infection Service, Public Health England, London, UK; Department of Ecology and Evolutionary Biology, University of California, Santa Cruz, California, USA

## Abstract

Quarantining close contacts of individuals infected with SARS-CoV-2 for 10 to 14 days is a key strategy in reducing transmission. However, quarantine requirements are often unpopular, with low adherence, especially when a large fraction of the population has been vaccinated. Daily contact testing (DCT), in which contacts are required to isolate only if they test positive, is an alternative to quarantine for mitigating the risk of transmission from traced contacts. In this study, we developed an integrated model of COVID-19 transmission dynamics and compared the strategies of quarantine and DCT with regard to reduction in transmission and social/economic costs (days of quarantine/self-isolation). Specifically, we compared 10-day quarantine to 7 days of self-testing using rapid lateral flow antigen tests, starting 3 days after exposure to a case. We modelled both incomplete adherence to quarantine and incomplete adherence to DCT. We found that DCT reduces transmission from contacts with similar effectiveness, at much lower social/economic costs, especially for highly vaccinated populations. The findings were robust across a spectrum of scenarios with varying assumptions on the speed of contact tracing, sensitivity of lateral flow antigen tests, adherence to quarantine and uptake of testing. Daily tests would also allow rapid initiation of a new round of tracing from infected contacts.

## Introduction

Widespread testing and contact tracing have formed two important pillars of the COVID-19 response worldwide, with variable success in their implementation. Contact tracing identifies and informs individuals who have had close contact with a case, and usually requires that such individuals quarantine (stay at home) to reduce their chance of infecting others. Several factors make contact tracing for SARS-CoV-2 challenging (Ferretti, Wymant, et al. 2020): individuals become infectious before symptoms appear (Tindale et al. 2020; He et al. 2020), and can be infectious without ever showing symptoms (Buitrago-Garcia et al. 2020; Lavezzo et al. 2020); also, few individuals cause a large fraction of infections, often in so-called super-spreader events (Endo et al. 2020; Sun et al. 2021). Contact tracing has proven highly effective if cases are detected early and contacts are quarantined effectively. Countries that have adopted rigorous strategies for quarantine after contact with a confirmed case and at arrival in the country have generally fared well (Patel and Sridhar 2020; Oliu-Barton et al. 2021). Countries without rigorous enforcement have often seen only incomplete adherence. For example, in a UK survey, only 11% of contacts of a confirmed COVID-19 case self-reported full adherence to a 14-day quarantine, 54% partial adherence and 35% no adherence (Smith et al. 2021).

An alternative to quarantining contacts is daily contact testing (DCT), in which contacts are required to self-isolate only if they test positive. Where sufficient testing capacity is available, DCT can reduce the immediate social/economic costs imposed on contacts by quarantine since uninfected individuals don’t have to quarantine. However, the reduction in transmission from contacts using DCT must be carefully evaluated, because DCT might not detect all infected contacts and failing to prevent transmission can impose greater social/economic costs in the longer term. The ability of DCT to reduce transmission depends on how infectious contacts are, how sensitive the tests used are in practice, and the relative timing of transmission and test results.

The gold standard for COVID-19 testing is PCR testing, as it can detect viral loads as low as 100 copies per mL and therefore can detect early infections. However, PCR tests also report positive results after individuals have ceased to be infectious (Alexandersen, Chamings, and Bhatta 2020). During the pandemic, other rapid tests with lower sensitivity have become available, e.g. loop-mediated isothermal amplification tests, transcription-mediated amplification tests, and lateral flow antigen (LFA) tests. All tests based on oropharyngeal swabs, including PCR tests, tend to be less sensitive when not performed by a trained individual (Lindner et al. 2021; Peto and UK COVID-19 Lateral Flow Oversight Team 2021; Stohr et al. 2021; Coste, Egli, and Greub 2021). Limitations resulting from partial swabbing, lower test sensitivity and incomplete adherence can be overcome by designing testing schemes that make the best use of all tests available.

LFA tests use antibodies to detect the presence of spike protein, a component of the viral envelope, with much less sensitivity than PCR tests: LFA tests detect positive samples with 10,000 or more virions per mL elution from an oropharyngeal swab. Policies for these tests must be designed according to their predictive values in different circumstances: if the purpose of testing is to find cases and promote self-isolation before transmission, coverage and availability of testing is far more important than sensitivity for control (Larremore et al. 2020), in line with the argument that the sensitivity of the overall testing strategy should be considered, not just the sensitivity of a single test within it (Mina, Parker, and Larremore 2020). A strong dependence of infectiousness on viral load is key for this argument, since this induces correlation between infectiousness and test sensitivity, and therefore predicts that less sensitive tests preferentially identify the individuals most infectious at the time of testing.

The challenge of assessing the usefulness of LFA tests is using the right parameters for the infection dynamics of COVID-19, taking into account the interdependence between viral load at different stages of disease, when individuals develop symptoms, and when they become infectious. Averages may not correctly account for potentially relevant variability between individuals. For example, early studies considered identical infectiousness and sensitivity curves for each individual (Grassly et al. 2020), whereas longitudinal studies on individuals (Kissler, Fauver, Mack, Olesen, et al. 2021; Jones et al. 2021) show that viral load trajectories have generally sharp peaks, occurring on different days after infection. Models which account for this covariance between viral load, infectiousness and test sensitivity suggest that LFA tests are more sensitive than earlier studies suggested (Larremore et al. 2020). The impact of LFA testing strategies on transmission using realistic parameters for viral load trajectories has been modelled in detail (Quilty et al. 2021) but only under the simplifying assumption of fixed infectiousness above a threshold in viral load.

In this study, we evaluated the effectiveness of daily LFA testing using a model of SARS-CoV-2 transmission dynamics that incorporates new information on the relationship between viral load and infectiousness (Lee et al. 2021). Our model is suitable for evaluating repeated testing strategies for different policy objectives, acknowledging the uncertainty in the relationship between viral load and infectiousness. We focused on the specific case of DCT. We explored under which circumstances DCT is more effective at reducing onward transmission than a 10-day quarantine with varying levels of incomplete adherence, taking into account the social/economic costs and the extent of vaccination.

## Results

### An integrated model for COVID-19 transmission dynamics

We developed a model describing viral load trajectories, onset of symptoms and individual infectiousness in time. We modelled the trajectory of viral loads in each individual as exponential growth until a peak, followed by exponential decline, with lognormal distributions describing variation among individuals for the rates of growth and decline, the initial value, and the peak value. We modelled infectiousness as a Hill function of viral load. For symptomatic cases, we modelled the time of onset of symptoms as a Gaussian-distributed delay relative to the peak of viral load. We included an additional exponential suppression of infectiousness after symptom onset, reflecting self-isolation or adoption of other precautionary behaviour.

This model takes into account several novel aspects including the individual viral load and transmission dynamics of COVID-19 and reproduced distributions of key biological parameters. In particular, we use data from individual viral load trajectories (Kissler, Fauver, Mack, Olesen, et al. 2021) which show a steeper growth phase than the viral load trajectories published previously, confirmed recently in a larger study by (Jones et al. 2021). We also use the viral load of the index case in modeling the probability of onward transmission, which reflects both direct measurements (Lee et al. 2021) and indirect information on the timing of transmission (Ferretti, Ledda, et al. 2020). Modeling the connection between viral load and infectiousness allows us to estimate how many onward transmissions a low-sensitivity test will miss compared to a high-sensitivity test.

Given the wide uncertainties in the shape of the viral load-infectiousness curve, we considered two different versions of the model: one uses the empirical curve found by Lee et al (Lee et al. 2021) from contact tracing data, and the other uses parameters inferred from the timing of exposure and transmission in case-contact pairs with partly known exposure and symptom dates. These correspond to two widely different infectiousness profiles, one with a gradual increase in infectiousness, the other with threshold-like behaviour at around 10^4^ viral copies/ml (Figure 1). Hence, consistency in outcomes between both versions of the model provides a strong check for the robustness of the results. Both models were calibrated to provide realistic incubation period distributions (Supplementary Figure 1), similar distributions of generation time for symptomatic individuals (Supplementary Figure 2) and time from onset of symptoms to transmission (Supplementary Figure 3). The viral load dynamics are assumed to be similar for all viral strains (Kissler, Fauver, Mack, Tai, et al. 2021; Ke et al. 2021).

**Figure 1:**
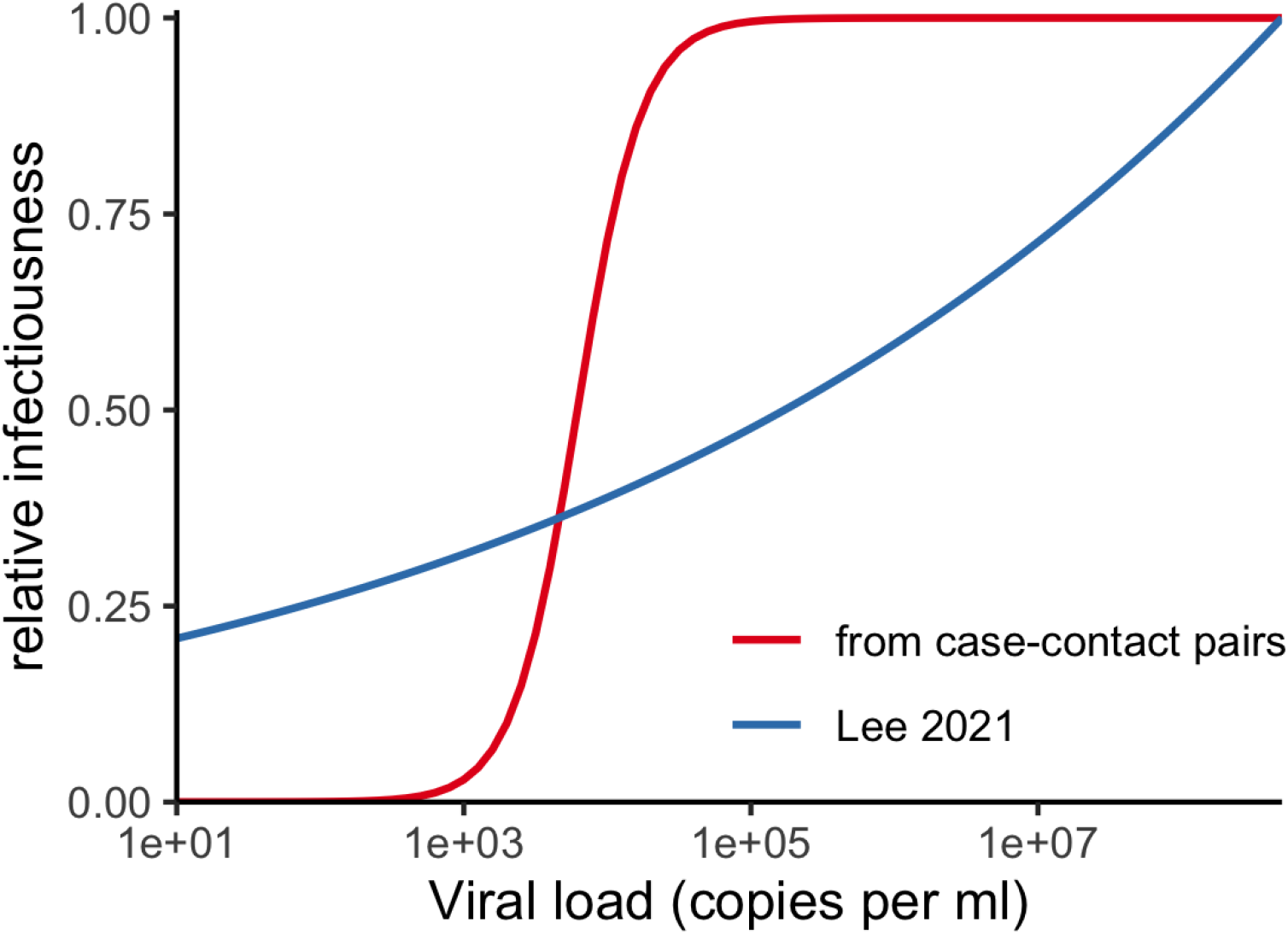
the relationship between viral load and infectiousness as inferred by Lee et al (Lee et al. 2021) shown in blue, and as inferred by Bayesian analysis of case-contact pairs, shown in red. Both curves are normalised with respect to the infectiousness corresponding to the maximum viral load considered in the analysis by Lee et al.

Infections in vaccinated individuals tend to cause fewer symptoms compared to unvaccinated individuals, and to have lower viral loads (Emary et al. 2021). Asymptomatic unvaccinated individuals also have reduced viral loads compared to mildly symptomatic individuals (as well as reduced symptoms, by definition) (Kissler, Fauver, Mack, Olesen, et al. 2021; Jones et al. 2021); we therefore approximated the viral load dynamics of vaccinated individuals with those previously measured for asymptomatic individuals. Vaccination also reduces the risk of getting infected (Emary et al. 2021). The precise reduction in risk varies with the viral strain, the type of vaccine, the vaccine recipient, and the time since vaccination; for simplicity we assumed a constant 75% reduction for the Delta variant, intermediate between the findings in (Lopez Bernal et al. 2021; Elliott et al. 2021).

### Sensitivity of single and repeated daily LFA tests

We compared a strategy of DCT using 7 days of LFA tests against a strategy of 10 days of quarantine, starting from the day the contact is notified, which we assume is 3 days after exposure. Durations were chosen based on a sensitivity analysis (see next section). The comparison depends on several factors. Since tests are self-administered, we included a 25% reduction in sensitivity relative to analytical sensitivity measured in ideal conditions (i.e. 25% false negative rate due to suboptimal testing, Supplementary Figure 4). We also included imperfect compliance with daily testing, assuming that contacts would miss any daily test at random with 20% probability, and would drop out of the testing regime altogether with a probability of 5% every day.

With these assumptions, the cumulative sensitivity of the test among all contacts found 3 days after exposure is shown in Figure 2, including the effects of the initial uptake, drop-out rate, and reduced sensitivity due to self testing. For one-off self-tests (orange dots), sensitivity starts as low as 50% and declines the later the test is taken. However, in the case of seven consecutive tests (black dots), overall sensitivity increases with every additional test and exceeds 95% by the seventh day. This shows that the cumulative sensitivity of repeated lateral flow testing is comparable to the one of a single clinically administered PCR test.

**Figure 2:**
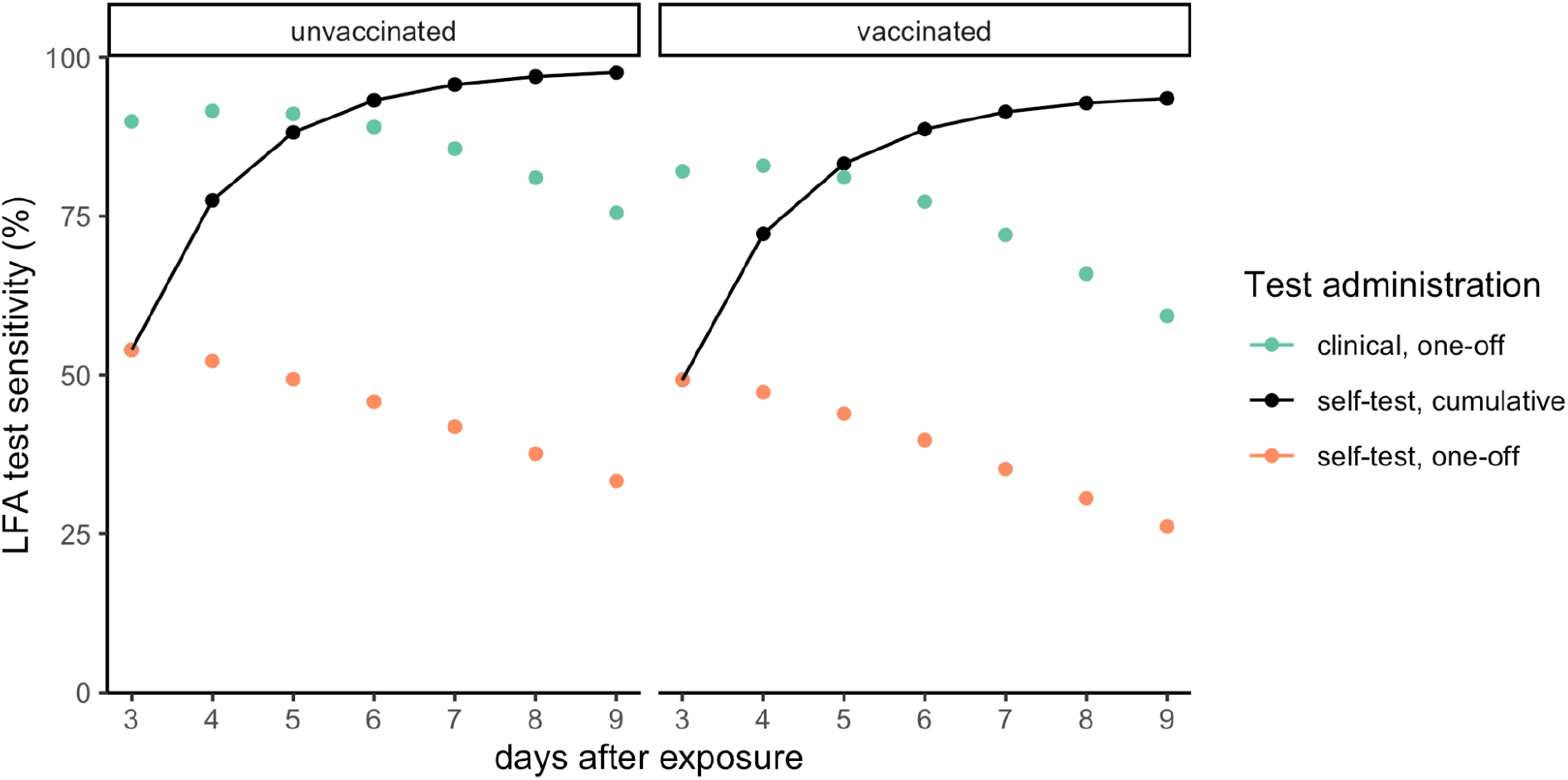
Repeated LFA testing has a sensitivity comparable to a single PCR test. LFA test sensitivity throughout the course of the infection, assuming that the first test is taken 3 days after exposure of the contact to their infector. The orange dots show the sensitivity of a one-off self-test, varying the day on which this is taken, including a 5% daily drop-out rate from testing. The black line shows the cumulative sensitivity of daily self-testing until that day. Both of these include a 25% reduction in sensitivity due to self-administration. As a comparison, the sensitivity of a one-off LFA test administered in a clinical setting (i.e. no reduction from self-administration and no drop-outs) is shown by green dots.

### Effectiveness of DCT is comparable to imperfect quarantine

At the time of writing in 2021, in the UK, contacts of cases are required to quarantine for 10 days. If this policy were followed faithfully by all such contacts, it would strongly reduce their further onward transmission. However, surveys suggest that adherence to quarantine may be low. For example, in the UK, one survey found self-reported adherence was 18% for self-isolation, and 11% for quarantine (Smith et al. 2021), while self-reported intent to adhere was 70%, and 65% for quarantine. Other surveys paint a more optimistic picture, with the COVID-19 Social Study reporting greater than 80% adherence (Fancourt et al. 2020) and experimental statistics from the Office of National Statistics suggest greater than 90% self-reported adherence (Brown 2021). We used an intermediate value between those reported by Smith et al. and Fancourt et al. as a reference (Wymant et al. 2021) (see Supplementary Methods). In this scenario, 75% of contacts adhere to quarantine, with an average 80% reduction in contact rates among contacts who adhere, and therefore an overall result of a 60% reduction in transmission during the prescribed quarantine period. However, since adherence to quarantine is uncertain and may decrease due to vaccinations and relaxation of COVID-19 related measures, we explore a wide range of adherence from 30% to 90%.

We also considered several behavioural factors for DCT, summarized here with our central assumptions:

- Initial uptake of daily testing (i.e. probability to collect the tests and start using them): 50%
- Probability of missing a test at random: 20%
- Daily drop-out rate from testing regime: 5%
- Effective reduction in contact rates during testing period, before a positive result or if no test is taken: 20%
- Effective reduction in contact rates after a positive result: 80%

We found that under these assumptions, DCT with 50% uptake is almost as effective in averting transmissions as quarantine with our central estimate of 75% adherence (Figures 3 and 4). This is true for both vaccinated and unvaccinated contacts. However, even with a relatively high 20% secondary attack rate (SAR, the probability of an individual being infected given that they have been notified), the social/economic costs (measured in days of quarantine/self-isolation) of the DCT strategy is considerably lower than the quarantine strategy, for both vaccinated and unvaccinated contacts. For lower SARs, corresponding to e.g. lower-risk exposures or higher vaccination uptake among contacts, the cost reduction would be still greater.

**Figure 3:**
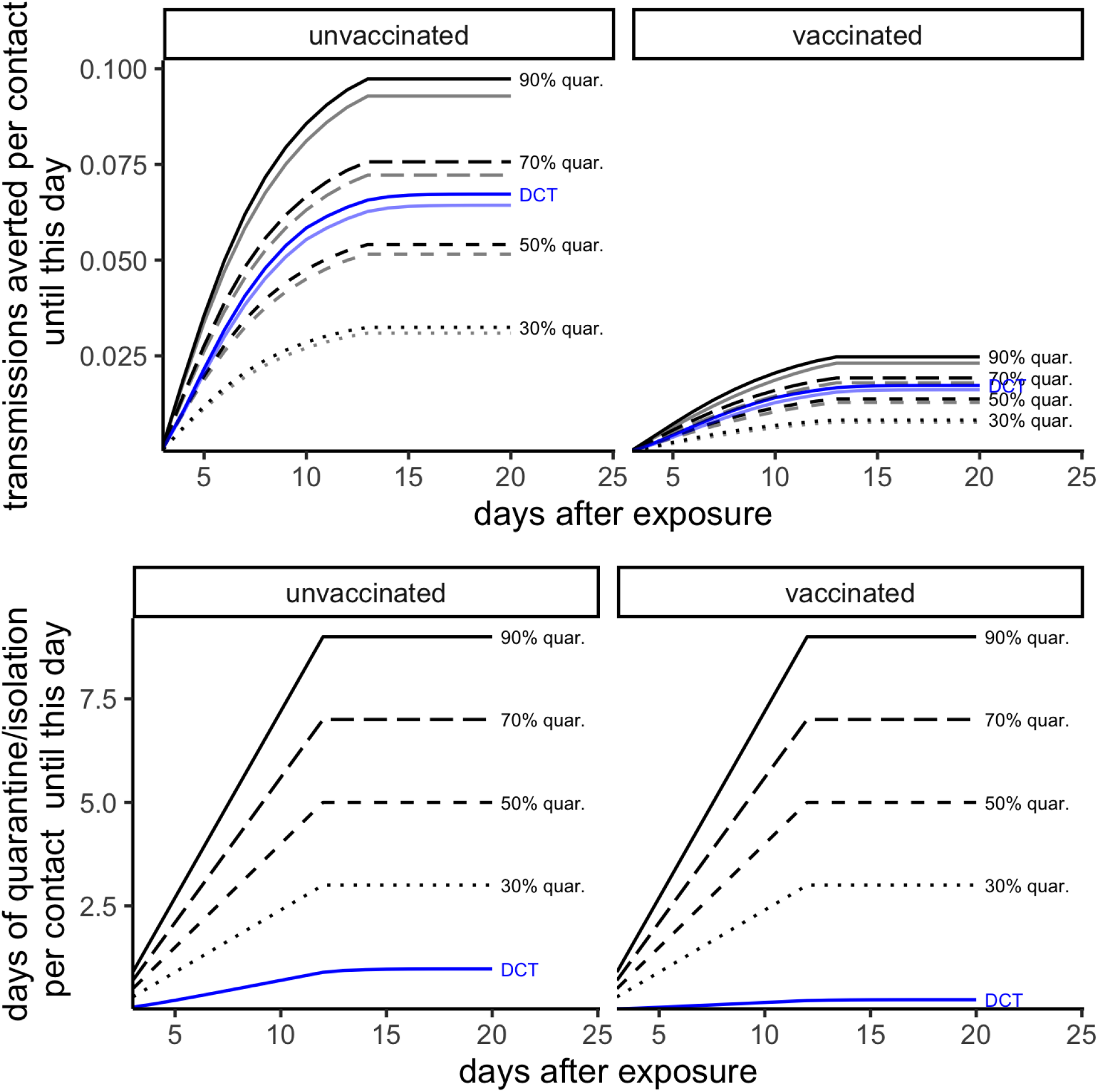
LFA testing reduces transmission over time as effectively as quarantine with intermediate compliance, and reduces social/economic costs. Top row: the cumulative number of transmissions averted on average from a contact-traced individual (who may or may not be infected) during the three weeks after exposure. Bottom row: the social/economic costs, measured as the average number of days of quarantine/self-isolation per traced individual. We assume that contacts are traced three days after their exposure to their infector and have a mean reproduction number of R=2 in the absence of contact tracing. The black lines show scenarios in which they follow quarantine imperfectly, with variable adherence (30, 50, 70, 90%) and no testing available. The blue line shows the alternative daily contact testing (DCT) scenario, in which contacts self-administer an LFA test every day, with greater (i.e. riskier) effective contact rates than the quarantine scenario before obtaining any positive test result, but lower (i.e. safer) effective contact rates after obtaining a positive result. Solid lines correspond to the inference of infectiousness from case-contact pairs, while semi-transparent lines correspond to the infectiousness from Lee et al.

**Figure 4:**
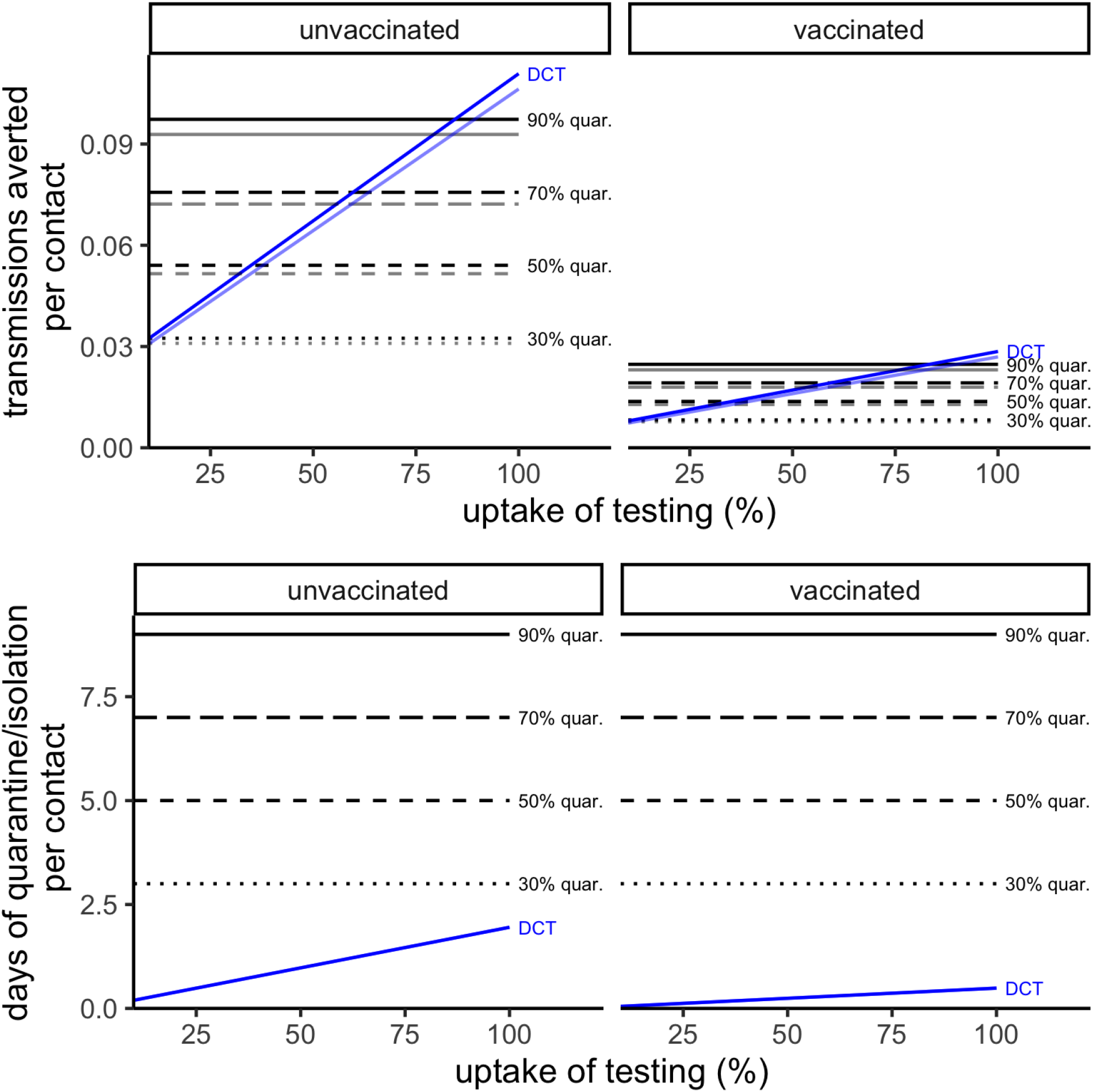
LFA testing reduces transmission more effectively than quarantine levels at reduced social/economic costs if testing levels are high. Top row: the total number of transmissions averted on average from a contact-traced individual over the course of the intervention. Bottom row: the social/economic costs, measured as the average number of days of quarantine/self-isolation per traced individual. The black lines show scenarios in which they follow quarantine imperfectly, with variable adherence (30, 50, 70, 90%) and no testing available. The blue line shows the alternative daily contact testing (DCT) scenario, in which contacts self-administer an LFA test every day, with greater (i.e. riskier) effective contact rates than the quarantine scenario before obtaining any positive test result, but lower (i.e. safer) effective contact rates after obtaining a positive result. The x-axis shows the effect of varying the initial uptake of tests from 30% to 100% (which applies to the DCT strategy only, with constant lines showing the quarantine strategy for comparison). Solid lines correspond to our Bayesian inference of infectiousness, while semi-transparent lines correspond to the infectiousness from Lee et al.

For the quarantine strategy, the socio-economic cost is the same for vaccinated and unvaccinated contacts. The benefit, in terms of reduced transmission, is greater for unvaccinated contacts than for vaccinated ones, but decreases more quickly as their adherence decreases. This is because unvaccinated individuals are more likely to be infected, and thus to infect others, so the benefit depends more sensitively on their adherence. For the DCT strategy, the social/economic cost is different for the two types of individuals: vaccinated individuals are less likely to be infected, to test positive, and to require isolation. By design, DCT allocates isolation in proportion to the contact’s risk of infecting others. This means the cost-benefit ratio is similar for the two types of individuals under DCT, whereas it is much higher for vaccinated than unvaccinated individuals under the quarantine strategy. For either type of individual, the cost-benefit ratio is much lower under DCT than the quarantine strategy (Supplementary Figure 5).

Solid and semi-transparent lines in Figure 3 and 4 show the same results, but with different models of how infectiousness depends on viral load (semi-transparent lines refer to the model based on the Lee 2021 result shown in Figure 1; solid ones to our new inference from case-contact pairs). The results are very similar, illustrating that the predictions are insensitive to the wide uncertainties in the shape of the viral load/infectiousness curve.

The effectiveness of DCT compared to quarantine also depends on how quickly contacts are traced, the rate of dropout from daily testing, the actual adherence to quarantine, etc. We explore realistic scenarios across a range of parameters (Supplementary Figure 6). In most reasonable scenarios modelled, 7-day DCT (blue line) with 50% uptake is broadly as effective in reducing onward transmission as a 10-day quarantine with survey-based adherence levels (black lines), and always has a much greater cost-benefit ratio. The results are robust to varying the duration of the DCT period between 5 and 10 days, with only marginal impact for vaccinated contacts and a more distinct improvement increasing the duration of DCT to 7-8 days for the unvaccinated contacts, but little improvement after that. Varying the duration of self-isolation/quarantine has a much greater impact on both transmissions averted and - naturally - days spent in isolation. The UK’s current policy of 10 days quarantine corresponds reasonably well to a knee in the curves for transmissions averted per unvaccinated contact for all but the 90% adherence to quarantine scenario, where a longer quarantine period may be deemed preferable.

The expected uptake of daily testing by contacts is especially uncertain, given the novelty of the policy, and hard to estimate before implementation. UK surveys suggest that about 46% of contacts would favour DCT while 41% would prefer to quarantine (Martin et al. 2021). Also, in a pilot study, about 62% of the contacts who did not already have access to testing accepted to join the pilot (Love et al. 2021). For this reason, we chose 50% uptake as our central scenario. However, the relative effectiveness of DCT and quarantine in reducing transmissions depends strongly on testing uptake and adherence to quarantine. To explore this dependence, Figure 4 illustrates the impact of varying uptake on both reduction in onward transmissions and days of quarantine/self-isolation. According to our model, an uptake of 25% for DCT would be equivalent to 50% adherence to quarantine in terms of reduction of onward transmissions, while an uptake of 80% would be equivalent to 90% adherence to quarantine.

## Discussion

We developed an integrated model of COVID-19 transmission and rapid LFA testing which incorporates key biological parameters consistent with the published literature. It builds on previous work (Larremore et al. 2020; Kissler, Fauver, Mack, Olesen, et al. 2021) and adds new insights about onward transmission as a function of viral load, variability among individuals in viral load trajectories, symptoms and vaccination status, and decreased sensitivity of self-testing compared to clinical testing, which are important aspects for assessing repeat self-application of low-sensitivity tests. LFA tests, or other COVID-19 rapid tests, could be useful in many scenarios that conduct regular, repeated testing (Mina, Parker, and Larremore 2020), e.g. for health care professionals, those caring for clinically vulnerable individuals, essential workers, contact bubbles within schools, and the general public. Daily testing has already been proposed as a viable strategy for several of these scenarios (Quilty et al. 2021) and has recently been shown to be an effective alternative to quarantine in schools (Leng et al. 2021; Young et al. 2021).

Here we focused on a daily contact testing (DCT) strategy, i.e. daily LFA testing as an alternative to quarantine of traced contacts. We estimated the reduction in onward transmission from individuals traced three days after exposure to a positive case by a DCT strategy, specifically of 7 daily LFA tests, and compared this to a strategy of 10-day quarantine. We found that, assuming intermediate adherence in both cases, the two strategies reduce onward transmission by a similar amount. However, the social/economic costs for DCT, measured in days of quarantine/self-isolation, were much lower. This result is robust across a variety of different scenarios based on varying assumptions. Shortening the duration of the testing period to 5 days does not significantly change the results. The advantage of DCT over quarantine increases with lower SAR (i.e. if the probability that contacts are actually infected is lower), and with lower adherence to quarantine. Increasing vaccination likely increases the advantage of DCT, since it reduces the probability of becoming infected, and may also shift public perception of risk and the importance of quarantine adherence.

One limitation of our results is our simple metric for the cost-benefit ratio, defined as the number of days of self-isolation or quarantine per transmission averted, for an average traced contact. When either top-down or voluntary control measures keep R(t) near 1 (Weitz et al. 2020), this metric describes efficiency in achieving this outcome (Petrie and Masel 2020). Translating the number of days spent in quarantine/self-isolation into specific social/economic costs will vary according to settings. The costs are particularly large, making DCT especially beneficial, in settings where individuals are unable to work from home, have no effective place to self-isolate, or struggle to live independently (Smith et al. 2021), as well as in settings where many people are repeatedly affected, for example where entire school year groups are sent home or workplaces closed (Leng et al. 2021). On the other hand, if reduced transmission tips the balance from an uncontrolled to a controlled epidemic, then even strategies with much larger costs for only slightly larger benefits (both defined as above) are preferred. For the present comparison between quarantine and DCT for traced contacts, the benefits were similar but costs were very different, simplifying the recommendation in favor of DCT in scenarios where the aim is epidemic mitigation, rather than elimination.

A further complication, given the low sensitivity of COVID-19 self-testing, is the need for clear communication. A negative result means the recipient is unlikely to be infectious at the time of the test, but their being infected is not ruled out, and they could become highly infectious in the near future after the test. This message should be emphasised when explaining DCT policies based on LFA tests to the general public.

We found that LFA testing compares more favourably to PCR testing than might be expected based on existing literature (Grassly et al. 2020; Hellewell et al. 2021; Quilty et al. 2021), despite a pessimistic assumption on the uptake of LFA tests (50%). This is partly due to the incorporation of individual viral load trajectories from (Kissler, Fauver, Mack, Olesen, et al. 2021). The faster growth phase estimated by this work means that individuals spend a shorter time in the window in which the viral load is sufficiently high to be detected by PCR, but not yet high enough to be detected by LFA tests. More important, however, is that we used new data on the relationship between viral load and infectiousness (Lee et al. 2021). According to these data, the fraction of transmissions that occur when the positive case has a high viral load is disproportionately higher than the fraction of transmissions that occur at lower viral load. This means that even though LFA tests miss more infections than PCR tests, the cases missed have lower viral loads and thus are those least likely to cause onward transmissions. Critical to the success we expect for LFA tests is their repeated use over a number of days, not as one-offs, which exponentially decreases the probability of an overall false negative result.

Replacing quarantine by DCT is not only expected to curb onward transmissions while reducing the negative impact of COVID-19 on the economy, but also has benefits via the more rapid identification of cases. Earlier diagnosis can lead to improved medical care, for example if contacts at high medical risk receive immediate prophylactic treatment (e.g mAbs). Further, once a contact is identified as a case a new round of contact tracing can be triggered (Bradshaw et al. 2021; Barrat et al. 2021). The contacts can infect others, either before being traced or in spite of it. With a quarantine strategy, individuals infected by a contact are only notified if and when the contact develops symptoms and obtains a PCR result. A DCT strategy, on the other hand, does not wait for that to happen: the next round of contact tracing can begin upon the first positive result of a rapid self-test, regardless of the extent or timing of symptoms, inclination to obtain a PCR test, and the speed of PCR testing. To maximise the benefit of such recursive tracing, we recommend integrating DCT with contact tracing apps where applicable. By adding a module to such apps that is able to read the result of LFA tests, positive test results can be acted upon immediately, reducing the time to recursive contact tracing and the prevention of further transmission.

## Methods

We jointly modelled the sensitivity of LFA tests and the infectiousness/transmission risk, considering how these vary as a function of viral load, both over the course of an infection and among individuals. The model was calibrated using step-wise Bayesian parameter inference coded in Stan (Carpenter et al. 2017), with parameters inferred from different data sources (Table 1). Details of the likelihood and inference can be found in Supplementary Methods.

**Table 1:** Datasets used to infer the biological parameters of the model.

|  | Source of information | Description | Reference |
| --- | --- | --- | --- |
| a) | Individual viral load trajectories | Longitudinal viral load measurements in individual patients | (Kissler, Fauver, Mack, Olesen, et al. 2021) |
| b) | Viral load trajectories and symptoms | Longitudinal viral load measurements with record of symptoms | (Hellewell et al. 2021) |
| c) | Incubation period distribution | Distribution of time intervals between infection and onset of symptoms | (McAloon et al. 2020) |
| d) | Timing of transmission | Distribution of time intervals from infection and onset of symptoms of person A to transmission to person B and onset of symptoms for B | (Ferretti, Ledda, et al. 2020) and datasets therein (Ferretti, Wymant, et al. 2020; Xia et al. 2020; He et al. 2020; Cheng et al. 2020) |
| e) | Viral load measurements and secondary attack rate | Viral load measurements with records of which fraction of household members were infected | (Lee et al. 2021) |
| f) | LFA test sensitivity | Dependence of test sensitivity on viral load in the sample (we consider Innova as an example) | (Peto and UK COVID-19 Lateral Flow Oversight Team 2021; Lee et al. 2021) |

The sensitivity of LFA tests was assumed to follow a logistic function of viral load, shown in Supplementary Figure 4 (Peto and UK COVID-19 Lateral Flow Oversight Team 2021).

Building on earlier modelling from Larremore et al. (Larremore et al. 2020) and especially the data and analysis from Kissler et al. (Kissler, Fauver, Mack, Olesen, et al. 2021), we modelled within-host viral load dynamics. Following (Kissler, Fauver, Mack, Olesen, et al. 2021), we modelled detectable Ct values as decreasing linearly until a nadir, then increasing linearly. Equivalently, given the linear relationship between Ct values and log viral load, detectable log viral load values were modelled as increasing linearly until a peak then decreasing linearly. Kissler et al. modelled the rates of increase and decrease, and the peak log viral load height, as normally distributed over the population of infected individuals; we modelled these same quantities as log-normally distributed (since all are non-negative, and the time to peak viral load is assumed linearly related to incubation time, which is log-normally distributed (McAloon et al. 2020)). These three parameters were independently estimated for asymptomatic individuals (no symptoms throughout the whole infection) and for symptomatic individuals (those who developed symptoms at some point, following a pre-symptomatic period). Inference for all parts of the model for viral load dynamics was implemented in Stan with the RStan interface (Guo et al. 2016), starting from code provided by Kissler et al., subsequently adapted for the present work. We re-fitted the model to obtain Bayesian posteriors for the underlying parameters, estimating them as the median of the marginal posterior distribution for each parameter.

After fixing the inferred distributions of peak viral load, growth and clearance rates, we used a similar approach to model the longitudinal data on Ct values from PCR testing and symptom tracking of UK healthcare workers from the SAFER study (Hellewell et al. 2021). We assumed a lognormally distributed incubation period using the meta-analysis result of (McAloon et al. 2020). The time of symptom onset was assumed to be normally distributed with respect to the time of peak viral load. Stan was then used to infer posterior distributions for the mean and variance of this normal distribution, and for the initial viral load at the time of infection. Point estimates of the parameters were obtained from the medians of the marginal posteriors as before. The SAFER study also provided an estimate of the baseline false negative rate of self-administered tests (see discussion in Supplementary).

A Hill functional form provides a natural relationship between viral load and infectiousness (Fraser et al. 2007). However, the relationship is not expected to be one-to-one throughout the whole course of infection. After the onset of symptoms, effective infectiousness – as measured by the hazard for transmission – is expected to be reduced due to biological effects and/or increased precautionary behaviour after symptom onset. We assumed that the infectiousness expected from a given viral load was reduced by a factor declining exponentially with time after onset of symptoms, reflecting a constant hazard to begin self-isolation or adopt other precautionary behaviour. We previously estimated the timing of effective transmissibility (Ferretti, Ledda, et al. 2020). Here we combined the datasets and the likelihood for the timing of transmission with the model for viral load and symptoms described above, in order to infer the parameters of the Hill-shaped relationship between viral load and infectiousness, as well as the behaviour-related rate of exponential decay after symptoms.

We inferred two different models for infectiousness as a function of viral load. For the first, we extracted the parameters of the Hill function directly from the logistic relationship between log viral load and attack rate across contacts, adjusted for covariates, as inferred empirically by (Lee et al. 2021) from a large dataset from contact tracing in the UK. For the second, we used the exponent of the Hill function from (Lee et al. 2021) as a weakly informative prior, and inferred all parameters. The likelihood for each of the steps and tables with the estimated parameters can be found in Supplementary Information.

## Data Availability

All the data used in this research is publicly available.

## Acknowledgements

We thank Stephen Kissler and Yonatan Grad for useful discussions and their help with the code from (Kissler, Fauver, Mack, Olesen, et al. 2021). This work was funded by a Li Ka Shing Foundation award to CF, and by research grant funding from the UK Department of Health and Social Care.

## Conflicts of interest

JB advised the UK government on the validation program for LFA tests. JM consults for WeHealth Solutions PBC, who distribute exposure notification solutions to Arizona and Bermuda.

## Notes

### Author Declarations

No IRB or ethics approval were necessary for this work.

## References

Alexandersen, Soren, Anthony Chamings, and Tarka Raj Bhatta. 2020. “SARS-CoV-2 Genomic and Subgenomic RNAs in Diagnostic Samples Are Not an Indicator of Active Replication.” Nature Communications 11 (1): 6059.

Barrat, A., C. Cattuto, M. Kivelä, S. Lehmann, and J. Saramäki. 2021. “Effect of Manual and Digital Contact Tracing on COVID-19 Outbreaks: A Study on Empirical Contact Data.” Journal of the Royal Society, Interface / the Royal Society 18 (178): 20201000.

Bradshaw, William J., Ethan C. Alley, Jonathan H. Huggins, Alun L. Lloyd, and Kevin M. Esvelt. 2021. “Bidirectional Contact Tracing Could Dramatically Improve COVID-19 Control.” Nature Communications 12 (1): 232.

Brown, Lynsey. 2021. “Coronavirus and Self-Isolation after Being in Contact with a Positive Case in England - Office for National Statistics.” Office for National Statistics. July 15, 2021. https://www.ons.gov.uk/peoplepopulationandcommunity/healthandsocialcare/conditionsanddiseases/bulletins/coronavirusandselfisolationafterbeingincontactwithapositivecaseinengland/28juneto3july2021.

Buitrago-Garcia, Diana, Dianne Egli-Gany, Michel J. Counotte, Stefanie Hossmann, Hira Imeri, Aziz Mert Ipekci, Georgia Salanti, and Nicola Low. 2020. “Occurrence and Transmission Potential of Asymptomatic and Presymptomatic SARS-CoV-2 Infections: A Living Systematic Review and Meta-Analysis.” PLoS Medicine 17 (9): e1003346.

Carpenter, Bob, Andrew Gelman, Matthew D. Hoffman, Daniel Lee, Ben Goodrich, Michael Betancourt, Marcus Brubaker, Jiqiang Guo, Peter Li, and Allen Riddell. 2017. “Stan: A Probabilistic Programming Language.” Journal of Statistical Software, Articles 76 (1): 1–32.

Cheng, Hao-Yuan, Shu-Wan Jian, Ding-Ping Liu, Ta-Chou Ng, Wan-Ting Huang, Hsien-Ho Lin, and Taiwan COVID-19 Outbreak Investigation Team. 2020. “Contact Tracing Assessment of COVID-19 Transmission Dynamics in Taiwan and Risk at Different Exposure Periods Before and After Symptom Onset.” JAMA Internal Medicine, May. https://doi.org/10.1001/jamainternmed.2020.2020.

Coste, Alix T., Adrian Egli, and Gilbert Greub. 2021. “Self-Testing for SARS-CoV-2: Importance of Lay Communication.” Swiss Medical Weekly 151 (May): w20526.

Elliott, P., D. Haw, H. Wang, O. Eales, C. Walters, K. Ainslie, C. Atchison, et al. 2021. “REACT-1 Round 13 Final Report: Exponential Growth, High Prevalence of SARS-CoV-2 and Vaccine Effectiveness Associated with Delta Variant in England during May to July 2021,” August. https://spiral.imperial.ac.uk/handle/10044/1/90800

Emary, Katherine R. W., Tanya Golubchik, Parvinder K. Aley, Cristina V. Ariani, Brian Angus, Sagida Bibi, Beth Blane, et al. 2021. “Efficacy of ChAdOx1 nCoV-19 (AZD1222) Vaccine against SARS-CoV-2 Variant of Concern 202012/01 (B.1.1.7): An Exploratory Analysis of a Randomised Controlled Trial.” The Lancet 397 (10282): 1351–62.

Endo, Akira, Centre for the Mathematical Modelling of Infectious Diseases COVID-19 Working Group, Sam Abbott, Adam J. Kucharski, and Sebastian Funk. 2020. “Estimating the Overdispersion in COVID-19 Transmission Using Outbreak Sizes Outside China.” Wellcome Open Research 5 (July): 67.

Fancourt, D., F. Bu, H. Wan Mak, and A. Steptoe. 2020. “Covid-19 Social Study.” Results Release 28.

Ferretti, Luca, A. Ledda, C. Wymant, L. Zhao, V. Ledda, L. Abeler-Dorner, and Others. 2020. “The Timing of COVID-19 Transmission.” MedRxiv, Publisher: Cold Spring Harbor Laboratory Press. https://www.medrxiv.org/content/10.1101/2020.09.04.20188516v2.

Ferretti, Luca, Chris Wymant, Michelle Kendall, Lele Zhao, Anel Nurtay, Lucie Abeler-Dörner, Michael Parker, David Bonsall, and Christophe Fraser. 2020. “Quantifying SARS-CoV-2 Transmission Suggests Epidemic Control with Digital Contact Tracing.” Science 368 (6491). https://doi.org/10.1126/science.abb6936.

Fraser Christophe, T. Déirdre Hollingsworth, Ruth Chapman, Frank de Wolf, and William P. Hanage. 2007. “Variation in HIV-1 Set-Point Viral Load: Epidemiological Analysis and an Evolutionary Hypothesis.” Proceedings of the National Academy of Sciences of the United States of America 104 (44): 17441–46.

Grassly, Nicholas C., Margarita Pons-Salort, Edward P. K. Parker, Peter J. White, Neil M. Ferguson, and Imperial College COVID-19 Response Team. 2020. “Comparison of Molecular Testing Strategies for COVID-19 Control: A Mathematical Modelling Study.” The Lancet Infectious Diseases 20 (12): 1381–89.

Guo, Jiqiang, D. Lee, K. Sakrejda, J. Gabry, B. Goodrich, J. De Guzman, E. Niebler, T. Heller, and J. Fletcher. 2016. “Rstan: R Interface to Stan.” R 534: 0–3.

Hellewell, Joel, The SAFER Investigators and Field Study Team, Timothy W. Russell, Rupert Beale, Gavin Kelly, Catherine Houlihan, Eleni Nastouli, Adam J. Kucharski, The Crick COVID-19 Consortium, and CMMID COVID-19 working group. 2021. “Estimating the Effectiveness of Routine Asymptomatic PCR Testing at Different Frequencies for the Detection of SARS-CoV-2 Infections.” BMC Medicine. https://doi.org/10.1186/s12916-021-01982-x.

He, Xi, Eric H. Y. Lau, Peng Wu, Xilong Deng, Jian Wang, Xinxin Hao, Yiu Chung Lau, et al. 2020. “Temporal Dynamics in Viral Shedding and Transmissibility of COVID-19.” Nature Medicine 26 (5): 672–75.

Jones, Terry C., Guido Biele, Barbara Mühlemann, Talitha Veith, Julia Schneider, Jörn Beheim-Schwarzbach, Tobias Bleicker, et al. 2021. “Estimating Infectiousness throughout SARS-CoV-2 Infection Course.” Science 373 (6551). https://doi.org/10.1126/science.abi5273.

Ke, Ruian, Pamela P. Martinez, Rebecca L. Smith, Laura L. Gibson, Agha Mirza, Madison Conte, Nicholas Gallagher, et al. 2021. “Daily Sampling of Early SARS-CoV-2 Infection Reveals Substantial Heterogeneity in Infectiousness.” medRxiv : The Preprint Server for Health Sciences, July. https://doi.org/10.1101/2021.07.12.21260208.

Kissler, Stephen M., Joseph R. Fauver, Christina Mack, Scott W. Olesen, Caroline Tai, Kristin Y. Shiue, Chaney C. Kalinich, et al. 2021. “Viral Dynamics of Acute SARS-CoV-2 Infection and Applications to Diagnostic and Public Health Strategies.” PLoS Biology 19 (7): e3001333.

Kissler, Stephen M., Joseph R. Fauver, Christina Mack, Caroline G. Tai, Mallery I. Breban, Anne E. Watkins, Radhika M. Samant, et al. 2021. “Densely Sampled Viral Trajectories for SARS-CoV-2 Variants Alpha (B.1.1.7) and Epsilon (B.1.429).” medRxiv, July, 2021.02.16.21251535.

Larremore, Daniel B., Bryan Wilder, Evan Lester, Soraya Shehata, James M. Burke, James A. Hay, Milind Tambe, Michael J. Mina, and Roy Parker. 2020. “Test Sensitivity Is Secondary to Frequency and Turnaround Time for COVID-19 Surveillance.” medRxiv : The Preprint Server for Health Sciences, June. https://doi.org/10.1101/2020.06.22.20136309.

Lavezzo, Enrico, Elisa Franchin, Constanze Ciavarella, Gina Cuomo-Dannenburg, Luisa Barzon, Claudia Del Vecchio, Lucia Rossi, et al. 2020. “Suppression of a SARS-CoV-2 Outbreak in the Italian Municipality of Vo’.” Nature 584 (7821): 425–29.

Lee, Lennard Y. W., Stefan Rozmanowski, Matthew Pang, Andre Charlett, Charlotte Anderson, Gareth J. Hughes, Matthew Barnard, et al. 2021. “SARS-CoV-2 Infectivity by Viral Load, S Gene Variants and Demographic Factors and the Utility of Lateral Flow Devices to Prevent Transmission.” Clinical Infectious Diseases: An Official Publication of the Infectious Diseases Society of America, May. https://doi.org/10.1093/cid/ciab421.

Leng, Trystan, Edward M. Hill, Alex Holmes, Emma Southall, Robin N. Thompson, Michael J. Tildesley, Matt J. Keeling, and Louise Dyson. 2021. “Quantifying within-School SARS-CoV-2 Transmission and the Impact of Lateral Flow Testing in Secondary Schools in England.” bioRxiv. medRxiv. https://doi.org/10.1101/2021.07.09.21260271.

Lindner, Andreas K., Olga Nikolai, Franka Kausch, Mia Wintel, Franziska Hommes, Maximilian Gertler Lisa, J. Krüger, et al. 2021. “Head-to-Head Comparison of SARS-CoV-2 Antigen-Detecting Rapid Test with Self-Collected Nasal Swab versus Professional-Collected Nasopharyngeal Swab.” The European Respiratory Journal: Official Journal of the European Society for Clinical Respiratory Physiology 57 (4). https://doi.org/10.1183/13993003.03961-2020.

Lopez Bernal Jamie, Nick Andrews, Charlotte Gower, Eileen Gallagher, Ruth Simmons, Simon Thelwall, Julia Stowe, et al. 2021. “Effectiveness of Covid-19 Vaccines against the B.1.617.2 (Delta) Variant.” The New England Journal of Medicine, July. https://doi.org/10.1056/NEJMoa2108891.

Love, Nicola, Derren Ready, Charlie Turner, Lucy Yardley, G. James Rubin, Susan Hopkins, and Isabel Oliver. 2021. “The Acceptability of Testing Contacts of Confirmed COVID-19 Cases Using Serial, Self-Administered Lateral Flow Devices as an Alternative to Self-Isolation.” bioRxiv. medRxiv. https://doi.org/10.1101/2021.03.23.21254168.

Martin, Alex F., Sarah Denford, Nicola Love, Derren Ready, Isabel Oliver,Richard Amlôt, G. James Rubin, and Lucy Yardley. 2021. “Engagement with Daily Testing instead of Self-Isolating in Contacts of Confirmed Cases of SARS-CoV-2.” BMC Public Health 21 (1): 1067.

McAloon, Conor, Áine Collins, Kevin Hunt, Ann Barber, Andrew W. Byrne, Francis Butler, Miriam Casey, et al. 2020. “Incubation Period of COVID-19: A Rapid Systematic Review and Meta-Analysis of Observational Research.” BMJ Open 10 (8): e039652.

Mina, Michael J., Roy Parker, and Daniel B. Larremore. 2020. “Rethinking Covid-19 Test Sensitivity - A Strategy for Containment.” The New England Journal of Medicine 383 (22): e120.

Oliu-Barton, Miquel, Bary S. R. Pradelski, Philippe Aghion, Patrick Artus, Ilona Kickbusch, Jeffrey V. Lazarus, Devi Sridhar, and Samantha Vanderslott. 2021. “SARS-CoV-2 Elimination, Not Mitigation, Creates Best Outcomes for Health, the Economy, and Civil Liberties.” The Lancet 397 (10291): 2234–36.

Patel, Jay, and Devi Sridhar. 2020. “We Should Learn from the Asia-Pacific Responses to COVID-19.” The Lancet Regional Health. Western Pacific 5 (December): 100062.

Peto, Tim, and UK COVID-19 Lateral Flow Oversight Team. 2021. “COVID-19: Rapid Antigen Detection for SARS-CoV-2 by Lateral Flow Assay: A National Systematic Evaluation of Sensitivity and Specificity for Mass-Testing.” EClinicalMedicine 36 (June): 100924.

Petrie, James, and Joanna Masel. 2020. “The Economic Value of Quarantine Is Higher at Lower Case Prevalence, with Quarantine Justified at Lower Risk of Infection.” bioRxiv. medRxiv. https://doi.org/10.1101/2020.11.24.20238204.

Quilty, Billy J., Samuel Clifford, Joel Hellewell, Timothy W. Russell, Adam J. Kucharski, Stefan Flasche, W. John Edmunds, and Centre for the Mathematical Modelling of Infectious Diseases COVID-19 working group. 2021. “Quarantine and Testing Strategies in Contact Tracing for SARS-CoV-2: A Modelling Study.” The Lancet. Public Health 6 (3): e175–83.

Smith, Louise E., Henry W. W. Potts, Richard Amlôt, Nicola T. Fear, Susan Michie, and G. James Rubin. 2021. “Adherence to the Test, Trace, and Isolate System in the UK: Results from 37 Nationally Representative Surveys.” BMJ 372 (March): n608.

Stohr, Jjjm, V. F. Zwart, G. Goderski, and A. Meijer. 2021. “Self-Testing for the Detection of SARS-CoV-2 Infection with Rapid Antigen Tests.” medRxiv. https://www.medrxiv.org/content/10.1101/2021.02.21.21252153v1.abstract

Sun, Kaiyuan, Wei Wang, Lidong Gao, Yan Wang, Kaiwei Luo, Lingshuang Ren, Zhifei Zhan, et al. 2021. “Transmission Heterogeneities, Kinetics, and Controllability of SARS-CoV-2.” Science 371 (6526). https://doi.org/10.1126/science.abe2424.

Tindale, Lauren C., Jessica E. Stockdale, Michelle Coombe, Emma S. Garlock, Wing Yin Venus Lau, Manu Saraswat, Louxin Zhang, Dongxuan Chen, Jacco Wallinga, and Caroline Colijn. 2020. “Evidence for Transmission of COVID-19 prior to Symptom Onset.” eLife 9 (June). https://doi.org/10.7554/eLife.57149.

Weitz, Joshua S., Sang Woo Park, Ceyhun Eksin, and Jonathan Dushoff. 2020. “Awareness-Driven Behavior Changes Can Shift the Shape of Epidemics Away from Peaks and toward Plateaus, Shoulders, and Oscillations.” Proceedings of the National Academy of Sciences. https://doi.org/10.1073/pnas.2009911117.

Wymant, Chris, Luca Ferretti, Daphne Tsallis, Marcos Charalambides, Lucie Abeler-Dörner, David Bonsall, Robert Hinch, et al. 2021. “The Epidemiological Impact of the NHS COVID-19 App.” Nature 594 (7863): 408–12.

Xia, Wei, Jiaqiang Liao, Chunhui Li, Yuanyuan Li, Xi Qian, Xiaojie Sun, Hongbo Xu, et al. 2020. “Transmission of Corona Virus Disease 2019 during the Incubation Period May Lead to a Quarantine Loophole.” Epidemiology. medRxiv. https://doi.org/10.1101/2020.03.06.20031955.

Young, Bernadette C., David W. Eyre, Saroj Kendrick, Chris White, Sylvester Smith, George Beveridge, Toby Nonnemacher, et al. 2021. “1 A Cluster Randomised Trial of the Impact of a Policy of Daily Testing for 2 Contacts of COVID-19 Cases on Attendance and COVID-19.” Preprint.

