## Supplementary Information for "Modelling the effectiveness and social costs of daily lateral flow antigen tests versus quarantine in preventing onward transmission of COVID-19 from traced contacts"

#### Supplementary Figures

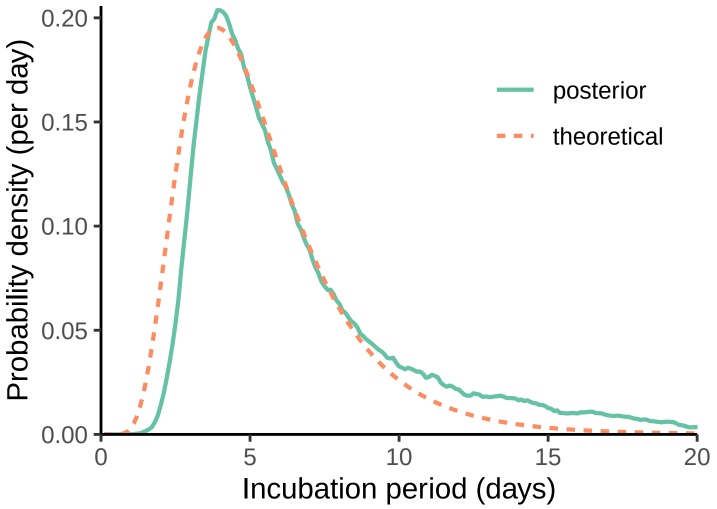


**Supplementary Figure** 1: Theoretical distribution of the incubation period used in the likelihood (orange dashed line), and simulated empirical distribution for the incubation period obtained from the inferred model (green solid line).

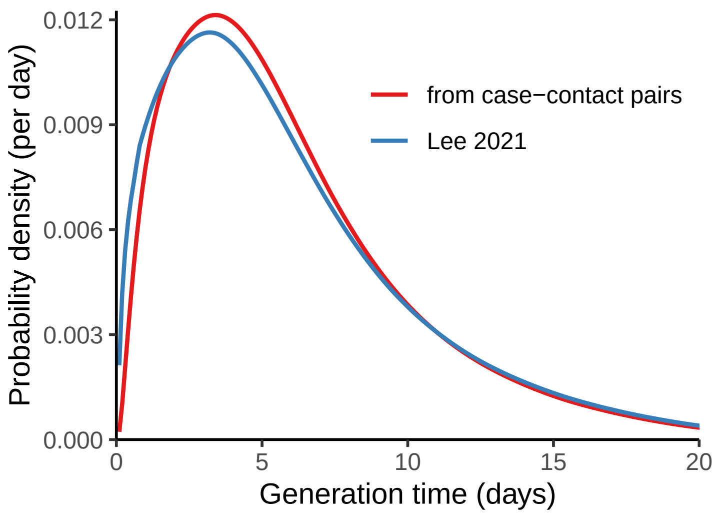


**Supplementary Figure** 2: Distribution of the generation time (the time from becoming infected oneself to infecting others), according to the two different relationships between infectiousness and viral load discussed in the text.

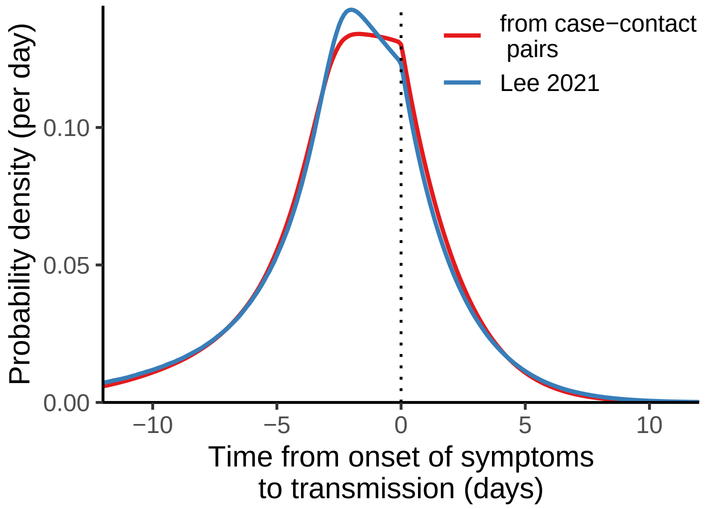


**Supplementary Figure** 3: Distribution of the time from onset of symptoms to transmission (TOST), according to the two different relationships between infectiousness and viral load discussed in the text.

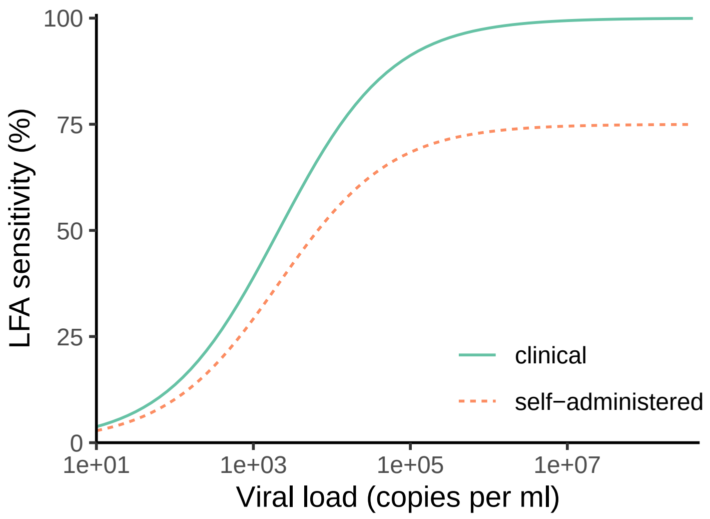


**Supplementary Figure** 4: Sensitivity of a LFA test depending on viral load. The dashed line shows the theoretical sensitivity from clinical samples in a laboratory setting; the continuous line illustrates the actual sensitivity that we considered for the model, accounting for self-administration. (Note that this sensitivity does not account for the probability of not taking some tests or dropping off the testing schedule altogether, which is considered separately in the model.)

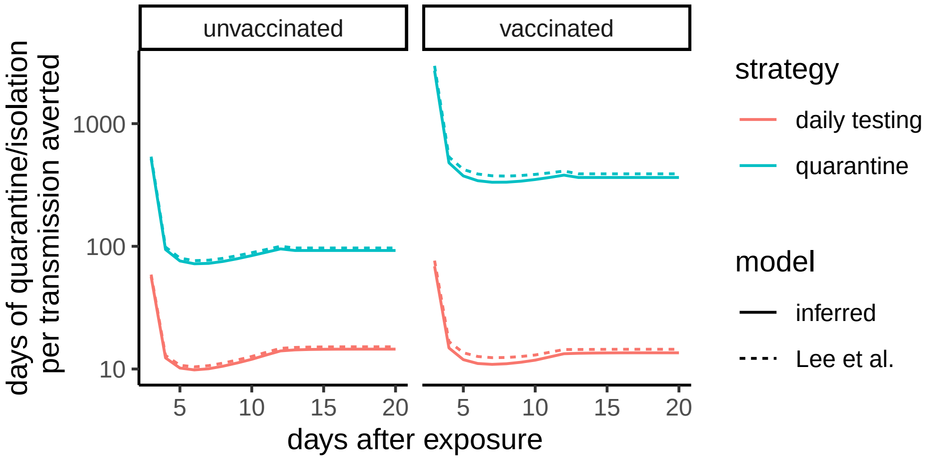


**Supplementary Figure** 5: Cost-benefit ratio for quarantine strategy and DCT strategy for vaccinated and unvaccinated contacts. The cost-benefit ratio is defined as the total days of quarantine or self-isolation per transmission averted. Results using the relationship between viral load and infectiousness inferred in this paper are shown as solid lines; results using the relationship inferred by Lee et al are shown as dashed lines.

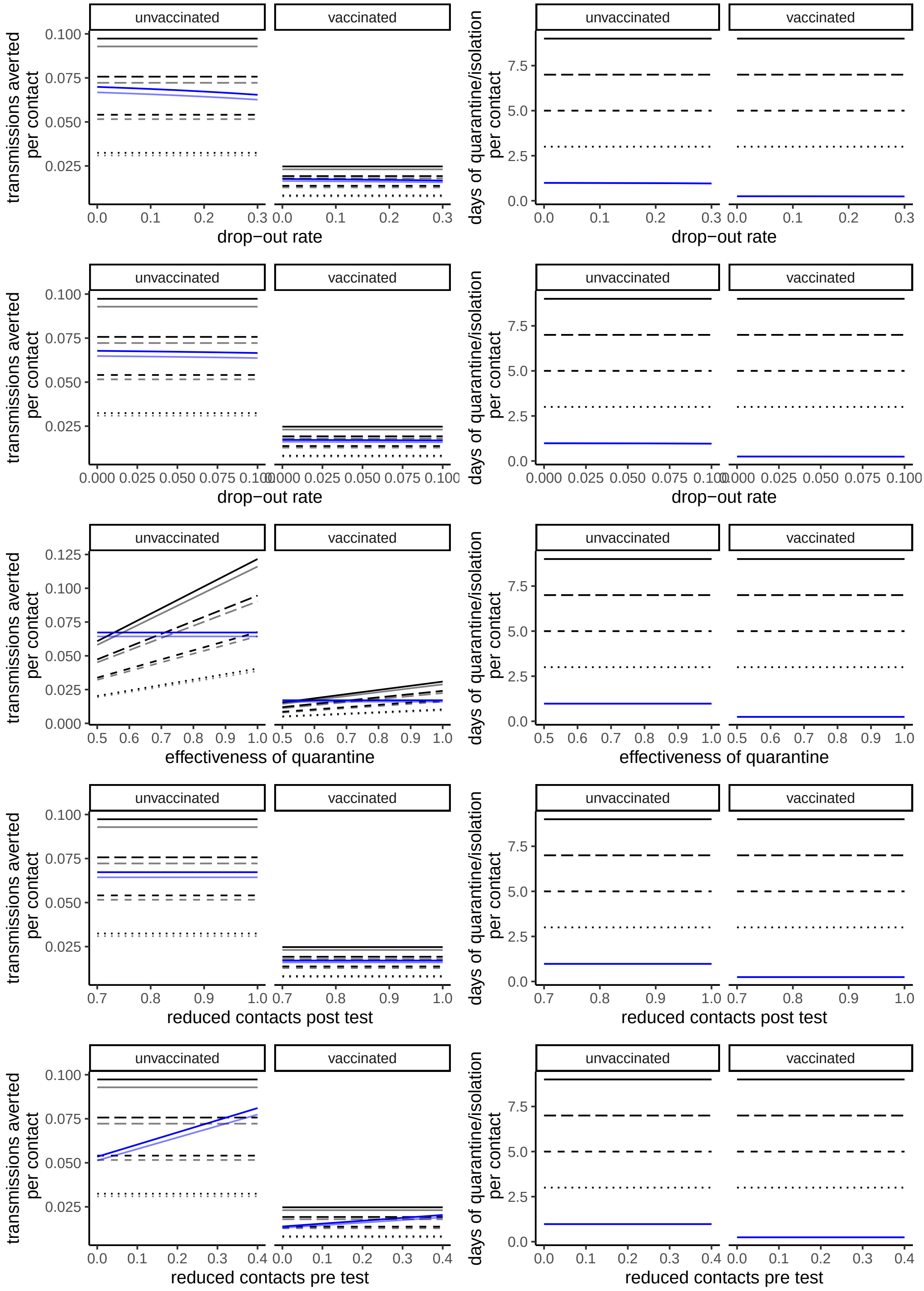
unvaccinated
vaccinated
transmissions averted per contact
days of quarantine/isolation per contact
0.000
0.025
0.050
0.075
0.100
0.125
0.0
2.5
5.0
7.5
drop−out rate
effectiveness of quarantine
reduced contacts post test
reduced contacts pre test

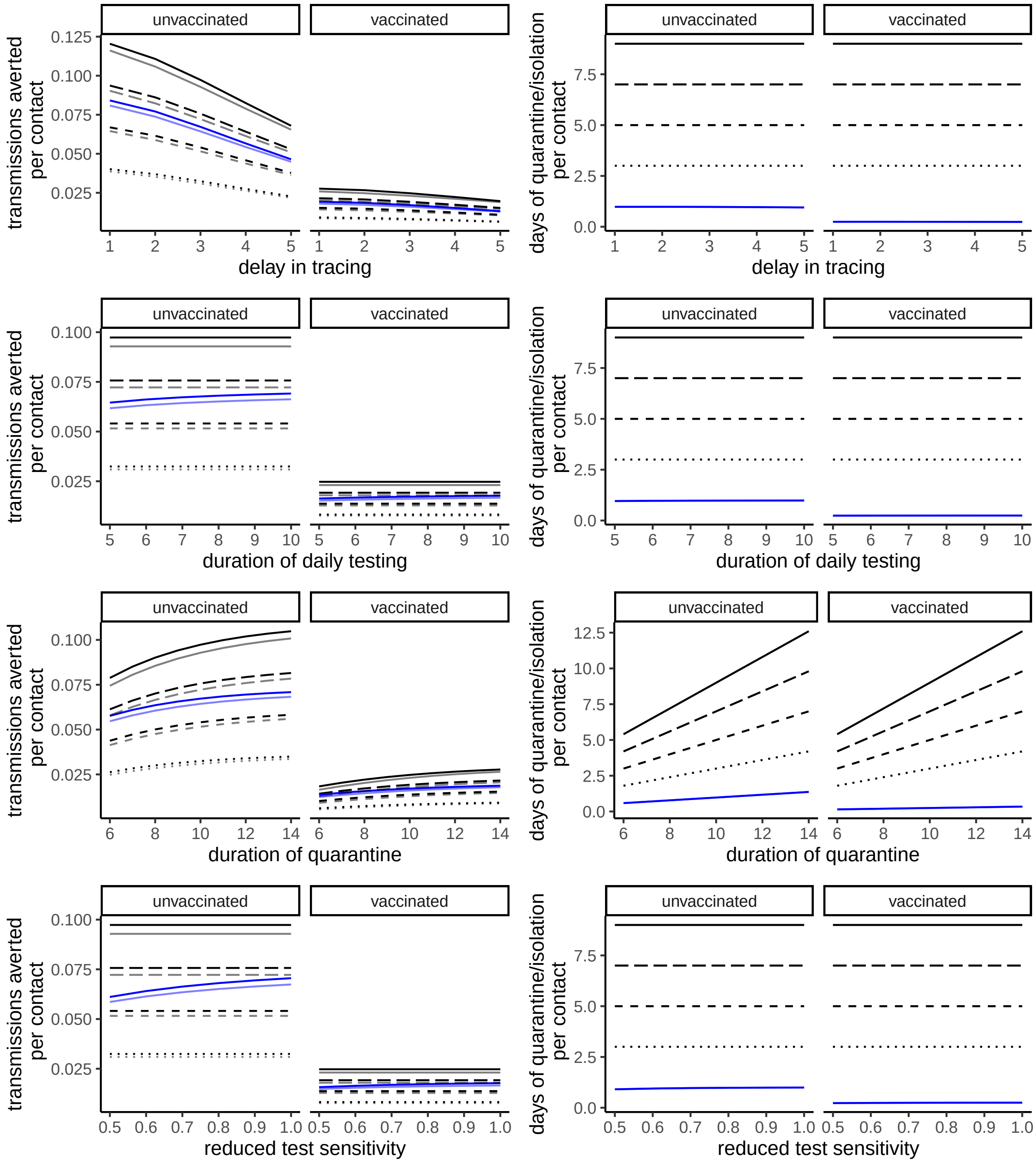


**Supplementary Figure** 6: Total number of transmissions averted by quarantine or DCT strategies for contact-traced individuals, and number of days of quarantine per traced individual. Quarantine is shown as black lines; daily testing is shown in blue. Solid lines correspond to inference of infectiousness from timing of transmission of case-contact pairs, semi-transparent lines to inference from Lee et al. Each row explores a different range for a given parameter with respect to the default scenario in Figure 3.

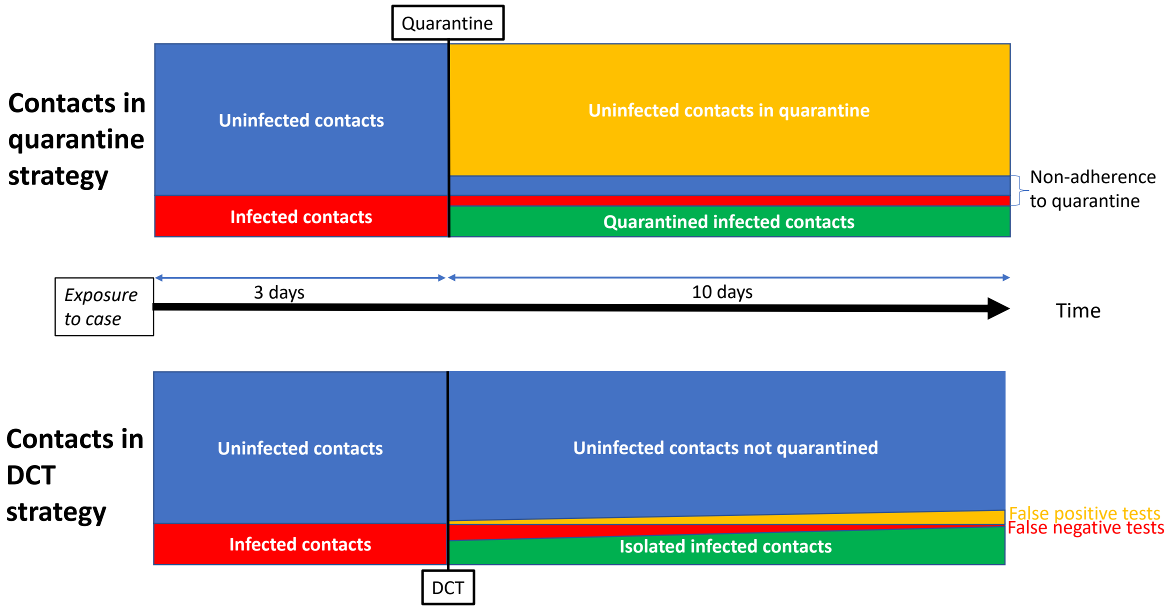


**Supplementary Figure 7**: Illustrative explanation of the lower cost/benefit ratio for DCT. The impact on transmission is given by the isolation of infected contacts (in green) from the general population, and it is comparable between quarantine and DCT with similar adherence. On the other hand, then number of uninfected contacts that are isolated (in yellow) ontributes to the difference in social costs between the two strategies, and it is disproportionately higher for quarantine.

### Supplementary Methods

#### Data sources

We bring together four main sources of data to parameterise the model:

- Serially sampled viral loads. [Kissler et al] have studied viral load trajectories (longitudinal Ct values) in serially sampled individuals in the NBA 'bubble' of players and support staff.
- Viral loads and symptoms. [Hellewell et al] provide serial positivity and Ct testing data with symptoms.
- Viral load/infectiousness curve. [Lee et al] fitted a logistic regression in terms of Ct values for the index case and other factors to the secondary attack rate in a large study on traced contacts from the UK.
- Timing of transmission. [Ferretti, Ledda et al] studied the time distribution between infection and/or onset of symptoms in the source cases, and transmission(s) to secondary cases, based on four datasets of case-contact pairs [Ferretti&Wymant et al, Xia et al, He et al, Cheng et al].

#### The model

Let $i$ index the different infected individuals.

##### Submodel 1

**Viral load trajectories** Time since infection is denoted $t$, and the viral load trajectory of individual $i$, denoted $V_i(t)$, is given by

$$V_i(t) = \begin{cases} V^0 \exp{(r_i t)}, & \text{if } t \leq t_i^P \\ V^0 \exp{(r_i t_i^P)} \exp{(-d_i(t - t_i^P))}, & \text{if } t > t_i^P \end{cases} \tag{1}$$

where

- $V^0$ is the viral load at the time of becoming infected for individual $i$,
- $r_i$ is the pre-peak exponential growth rate of viral load for individual $i$,
- $t_i^P$ is the time from becoming infected to peak viral load for individual $i$,
- $-d_i$ is the post-peak exponential 'growth' rate of viral load for individual $i$ (with $d_i > 0$ such that this describes exponential decline),

We re-parameterize this model by introducing a parameter for the peak viral load $V_i^P = V_i(t_i^P)$ and then noting that $t_i^P = (\log V_i^P - \log V_i^0)/r_i$.

We assume that all parameters $V_i^P, r_i, d_i$ are lognormally distributed with log-scale means $\mu^s_{\log V^P}$, $\mu^s_{\log r}$, $\mu^s_{\log d}$ for symptomatic individuals and $\mu^a_{\log V^P}$, $\mu^a_{\log r}$, $\mu^a_{\log d}$ for asymptomatic ones, and log-scale standard deviations $\sigma_{\log V^P}$, $\sigma_{\log r}$, $\sigma_{\log d}$. All distributions are truncated within realistic ranges to $r_i > \log(\mu_{V^P}/V_{lod})/15$, $d_i > \log(\mu_{V^P}/V_{lod})/30$, were $V_{lod}$ corresponds to the limit of detection, i.e. we assume that the visible growth and clearance phases cannot typically last more than 15 and 30 days respectively.

**Observational model** The rest of the model and the corresponding likelihood (dealing with the relative position of the peak, false negative and false positive rates, uncertainties in the measured Ct values vs the viral load trajectory) is identical to [Kissler et al].

Total viral load is not observed, rather viral load is assessed by quantitative polymerase chain reaction (qPCR) from a specimen obtained from the respiratory tract. The result is reported by the number of PCR cycles done before the virus is detected, the cycle threshold (Ct) value. The Ct value is specific to the method of testing, denoted $M$. $M$ includes details of the type of sample collected (e.g. nasopharygeal and oral swabs, saliva samples), methods of swabbing, and details of the laboratory protocol. The lower the Ct value, the higher the viral load. There is a limit of detection $\text{lod}_M$, that is specific to the testing method $M$. The expected Ct value is related to viral loads by the transform

$$E(Ct) = k_M - \log(VL)/e_M \tag{2}$$

where $k_M$ and $e_M$ are constants that are specific to the testing method $M$. Following [Kissler et al], we model the observed Ct value by

$$\Delta Ct \sim \lambda \text{ Normal}(E[\Delta Ct], \sigma_M) + (1-\lambda) \text{ Exponential}(\log 10)]_0^{\text{lod}} \tag{3}$$

where $\Delta Ct = \text{lod} - Ct$, $1 - \lambda = 0.01$ is the probability of a false positive result, and the right bracket indicates that the distribution is right truncated at lod.

Consider a time series of observed Ct values for individual $i$. This observation model defines a likelihood, subject to the unknown time of infection of individual $i$, $t_i^I$, which is the final parameter of the model.

Hierarchical Bayesian inference in Stan of the parameters of this submodel (except $V_i^0$, which cannot be inferred from pure longitudinal Ct information) results in the following estimates (with viral loads measured in Ct units, calibrated as $\log_{10} V = 2.658 + 0.277 \times Ct$):

| | mean | $se_{mean}$ | sd | 2.5% | 25% | 50% | 75% | 97.5% | $n_{eff}$ | $\hat{R}$ |
|---|---|---|---|---|---|---|---|---|---|---|
| $\mu^s_{\log VP}$ | 16.03 | 0.03 | 1.66 | 12.81 | 14.91 | 16.01 | 17.13 | 19.33 | 3555.27 | 1.00 |
| $\mu^s_r$ | 5.90 | 0.03 | 1.74 | 3.11 | 4.63 | 5.68 | 6.95 | 9.82 | 4081.83 | 1.00 |
| $\mu^s_d$ | -1.46 | 0.00 | 0.19 | -1.88 | -1.57 | -1.45 | -1.32 | -1.11 | 2342.91 | 1.00 |
| $\mu^a_{\log VP}$ | 12.98 | 0.03 | 1.13 | 10.78 | 12.22 | 12.95 | 13.74 | 15.22 | 1778.48 | 1.01 |
| $\mu^a_r$ | 4.32 | 0.02 | 1.09 | 2.65 | 3.55 | 4.18 | 4.91 | 6.92 | 2073.62 | 1.00 |
| $\mu^a_d$ | -1.60 | 0.01 | 0.19 | -2.02 | -1.72 | -1.59 | -1.46 | -1.26 | 967.87 | 1.01 |
| $\sigma_{\log V}$ | 4.10 | 0.01 | 0.68 | 2.87 | 3.63 | 4.05 | 4.52 | 5.54 | 2770.82 | 1.00 |
| $\sigma_{\log r}$ | 0.95 | 0.01 | 0.20 | 0.63 | 0.81 | 0.93 | 1.06 | 1.39 | 1271.81 | 1.01 |
| $\sigma_{\log d}$ | 0.24 | 0.01 | 0.09 | 0.06 | 0.18 | 0.24 | 0.30 | 0.44 | 330.38 | 1.02 |

##### Submodel 2

**Incubation and onset of symptoms** Denote by $t_i^S$ the onset of symptoms in individual $i$, defined only if individual $i$ develops symptoms at some point (i.e. is not asymptomatic). We assume that the onset of symptoms is driven by the peak of the viral load dynamics, and occurs at a mean time $\mu_{p-S}$ after the peak of viral load (or before, if $\mu_{p-S} < 0$). We model the self-reported onset of symptoms by a (truncated) normal distribution,

$$t_i^S \sim \text{Normal}(t_i^P + \mu_{p-S}, \sigma^2_{p-S}) \tag{4}$$

with the truncation corresponding to the condition $t_i^S > 0$ (i.e. symptoms shouldn't precede infection).

**Observational model** We apply the above viral load model to the data by [Hellewell et al] that include reported symptomatic/asymptomatic state along with test results. We exclude all observations from individuals without reported symptoms. Since Ct values from this study are not calibrated, the relation between Ct values and log viral load is modelled as linear $Ct = c_0 + c_1 \log(V)$ with parameters $c_0, c_1$ to be inferred.

The incubation period distribution $\iota(t)$ is assumed to be known (since it is derived from line lists of large numbers of individuals) and taken from the meta-analysis of [McAloon et al] with Ma et al. excluded due to possible bias. Namely, the incubation time $t_i^S$ is assumed to be lognormally distributed with $\mu_i = 1.58$, $\sigma_i = 0.47$. All times of infection and symptom onset have flat priors.

The likelihood for the viral load trajectories is similar to the previous one, except for the addition of a false negative rate $\lambda_{FNR}$ included in the model to account for self-testing, i.e. we model the observed Ct value by

$$\Delta Ct \sim \begin{cases} \lambda \text{Normal}(E[\Delta Ct], \sigma_M) + (1-\lambda)\ \text{Exponential}(\log 10)]_0^{\text{lod}} & \text{with prob } 1-\lambda_{FNR} \\ 0 & \text{with prob } \lambda_{FNR} \end{cases} \tag{5}$$

In submodel 2 we infer *de novo* all parameters, except the hyperpriors inferred in the previous submodel.

Hierarchical Bayesian inference in Stan of the parameters of this submodel and $V^0$ results in the following estimates:

| | mean | $se_{mean}$ | sd | 2.5% | 25% | 50% | 75% | 97.5% | $n_{eff}$ | $\hat{R}$ |
|---|---|---|---|---|---|---|---|---|---|---|
| $\log V^0$ | -1.21 | 0.02 | 1.19 | -4.37 | -1.70 | -0.85 | -0.34 | -0.02 | 3302.96 | 1.00 |
| $\mu_{p-S}$ | 2.50 | 0.01 | 0.45 | 1.66 | 2.20 | 2.49 | 2.79 | 3.39 | 1171.01 | 1.01 |
| $\sigma_{p-S}$ | 0.60 | 0.02 | 0.50 | 0.03 | 0.23 | 0.49 | 0.84 | 1.83 | 669.22 | 1.01 |
| $\lambda_{FNR}$ | 0.16 | 0.00 | 0.06 | 0.05 | 0.12 | 0.15 | 0.20 | 0.29 | 2652.82 | 1.01 |

Note that the estimate for $\lambda_{FNR}$ (16%) is lower than the FNR at peak sensitivity found in the work of [Hellewell et al] (23%). The first value is more likely to represent the true FNR due to self-administration by trained individuals. However, for the purpose of this model, we prefer to use a more conservative value and approximate the assumed FNR as 25%.

##### Submodel 3

**Viral infectiousness and transmissibility** We assume that infectiousness is a function of viral load and of time since symptoms. It is a monotonically increasing function of viral load, since high viral load should correlate with high viral shedding and higher transmission risk. We model it as a Hill function. We include also an exponential suppression of the infectiousness after onset of symptoms, due to behavioural effects (self-isolation and distancing).

$$\beta(V) = \beta_{max} \frac{V^\gamma}{V^\gamma + V_{50}^\gamma} \exp\left(-b\left[t - t_i^S\right]_+\right) \tag{6}$$

The parameters $\beta_{max}$, $\gamma$, $V_{50}$, $b$ are not assumed to vary between individuals, i.e. heterogeneity in the infectiousness between individuals is modelled as arising only though viral load and time since infection.

**Observational model** Observations of transmission events typically correspond to the exposure interval during which $i$ could have been infected, $[t_{i,\min}^E, t_{i,\max}^E]$ and the onset of symptoms

$T_i^S$ each individual $i = 1$ or $i = 2$ in a linked transmission pair. For small $\beta$, the likelihood of this observation is well approximated by

$$\int_{t^E_{1,\min}}^{t^E_{1,\max}} dt_1^E \int_{t^E_{2,\min}}^{t^E_{2,\max}} dt_2^E \iota(t_1^S - t_1^E)\beta(V_1(t_2^E), t_2^E)\iota(t_2^S - t_2^E) \tag{7}$$

Contact events without transmission correspond to the likelihood:

$$\int_{t^E_{1,\min}}^{t^E_{1,\max}} dt_1^E \iota(t_1^S - t_1^E) \exp\left[-\int_{t^E_{2,\min}}^{t^E_{2,\max}} dt_2^E \beta(V_1(t_2^E), t_2^E)\right] \tag{8}$$

Bayesian inference in Stan of all parameters of this submodel, considering a mild prior on $\gamma \sim \text{lognormal}(\log 0.092, \log 10)$ based on [Lee et al], results in the following estimates:

| | mean | $se_{mean}$ | sd | 2.5% | 25% | 50% | 75% | 97.5% | $n_{eff}$ | $\hat{R}$ |
|---|---|---|---|---|---|---|---|---|---|---|
| $\log V_{50}$ | 8.72 | 0.02 | 0.56 | 7.59 | 8.35 | 8.73 | 9.10 | 9.78 | 499.23 | 1.00 |
| $\gamma$ | 1.99 | 0.04 | 0.58 | 1.06 | 1.60 | 1.93 | 2.31 | 3.38 | 245.88 | 1.02 |
| $b$ | 0.40 | 0.00 | 0.07 | 0.27 | 0.35 | 0.39 | 0.44 | 0.54 | 484.34 | 1.01 |

On the other hand, assuming the values $\gamma = 0.092$ obtained from a direct fit of the nonlinear inference from [Lee et al] and $\log V_{50}$ large $(= 4\mu^s_{\log VP})$, Bayesian inference in Stan of the remaining parameters of this submodel results in the following estimate:

| | mean | $se_{mean}$ | sd | 2.5% | 25% | 50% | 75% | 97.5% | $n_{eff}$ | $\hat{R}$ |
|---|---|---|---|---|---|---|---|---|---|---|
| $b$ | 0.38 | 0.00 | 0.07 | 0.26 | 0.33 | 0.37 | 0.42 | 0.52 | 549.00 | 1.00 |

#### Modelling DCT and quarantine

Average reduction in contacts during quarantine was estimated in [Wymant&Ferretti et al] by considering an intermediate scenario assuming that approximately 45% of traced contacts quarantine perfectly (100% reduction in transmission), 30% of traced contacts quarantine imperfectly with 50% reduction in transmission, and 25% of traced contacts do not quarantine at all (0% reduction in transmission). This corresponds to an adherence of 45%+30%=75% to quarantine, with an average effectiveness of (100%·45%+50%30%)/75%=80%. We consider this scenario (75% adherence, 80% effectiveness in reducing contacts when quarantining) as our central scenario, but vary the adherence from 30% to 90% to acknowledge both uncertainties and potential changes in time in adherence, which also could differ between vaccinated and unvaccinated contacts.

The daily sensitivity of LFA tests is modelled with the self-administered sensitivity curve from Supplementary Figure 2 (approximately valid for Innova) and reduced by several behavioural factors, especially the daily probability of missing a test $p_{miss}$ and the daily probability of dropping out altogether $p_{drop}$. Hence, daily sensitivity on day $t$ post tracing is modulated by the behavioural factor $(1-p_{miss})*(1-p_{drop})^{t-1}$ and by the clinical sensitivity (which depends on the viral load that day).

The difference in cost-benefit between DCT and quarantine is driven by two epidemiological parameters: the $SAR$ among contacts of unvaccinated index cases, and the vaccine protection against infection ($VPI$). The social/economic cost of the quarantine strategy is independent of these parameters, but the benefit in terms of the absolute reduction in transmissions is proportional to the $SAR$ for contacts of unvaccinated cases and to $SAR(1 - VPI)$ for contacts of vaccinated cases. Current variants have a relatively low $SAR$ [Lee et al. 2021] and a high $VPI$ [Hall et al. 2021], here assumed to be $SAR = 0.2$ and $VPI = 0.75$, leading therefore to a high cost per transmission averted, especially for vaccinated individuals. On the other hand, both the cost and the benefit of DCT have the same dependence on $SAR$ and $VPI$ as for quarantine, i.e they are both proportional to the $SAR$ for contacts of unvaccinated cases and to $SAR(1 - VPI)$ for contacts of vaccinated cases. For this reason, DCT has a much greater cost-benefit ratio than the quarantine strategy, especially for vaccinated contacts and for contacts of vaccinated individuals. Since the difference in cost-benefit ratio between the two strategies depends on the $SAR$, it could also vary depending on the infectiousness of the circulating viral variants, and on other interventions that affect how likely infection is to occur given close contact (for example masks, the movement of social mixing outdoors, and increasing ventilation indoors).
